# COVID-19 vaccine hesitancy worldwide: a systematic review of vaccine acceptance rates

**DOI:** 10.1101/2020.12.28.20248950

**Authors:** Malik Sallam

## Abstract

**Introduction:** Utility of vaccine campaigns to control coronavirus disease 2019 (COVID-19) is not merely dependent on vaccine efficacy and safety. Vaccine acceptance among the general public and the healthcare workers, appears to have a decisive role for successful control of the pandemic.

**Aim:** To provide an up-to-date assessment of COVID-19 vaccination acceptance rates worldwide.

**Methods:** A systematic search of the peer-reviewed English survey literature indexed in PubMed was done on December 25, 2020. Results from 30 studies, met the inclusion criteria and formed the basis for final COVID-19 vaccine acceptance estimates. Results of an additional recent survey from Jordan and Kuwait was considered in this review as well.

**Results:** Survey studies on COVID-19 vaccine acceptance rates were found from 33 different countries. Among adults representing the general public, the highest COVID-19 vaccine acceptance rates were found in Ecuador (97.0%), Malaysia (94.3%), Indonesia (93.3%) and China (91.3%). On the other hand, the lowest COVID-19 vaccine acceptance rates were found in Kuwait (23.6%), Jordan (28.4%), Italy (53.7), Russia (54.9%), Poland (56.3%), US (56.9%), and France (58.9%). Only eight surveys among healthcare workers (doctors, nurses) were found, with vaccine acceptance rates ranging from 27.7% in the Democratic Republic of the Congo to 78.1% in Israel. In a majority of survey studies among the general public (62%), the acceptance of COVID-19 vaccination showed a level of ≥ 70%.

**Conclusions:** Low rates of COVID-19 vaccine acceptance were reported in the Middle East, Russia, Africa and several European countries. This could represent a major problem in the global efforts that aim to control the current COVID-19 pandemic. More studies are recommended to address the scope of COVID-19 vaccine hesitancy. Such studies are particularly needed in the Middle East Africa, Eastern Europe, Central Asia, Middle and Latin America.

**Graphical Abstract:** 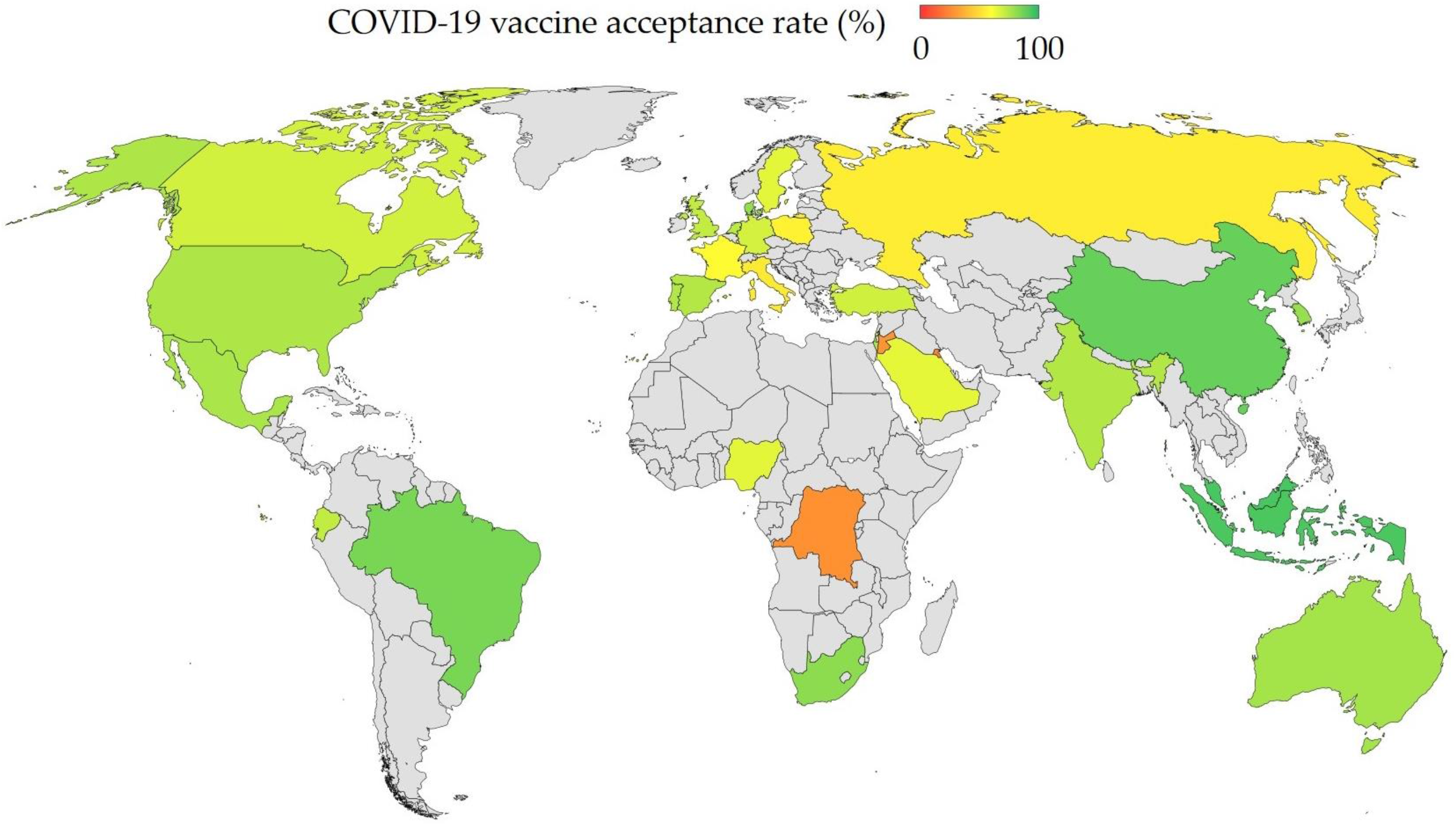

**COVID-19 vaccine acceptance rates worldwide:** For countries with more than one survey study, the vaccine acceptance rate of the latest survey was used in this graph. The estimates were also based on studies from the general population, except in the following cases were no studies from the general public were found (Australia: parents/guardians; DRC: healthcare workers; Hong Kong: healthcare workers; Malta: healthcare workers).

## Introduction

Based on Strategic Advisory Group of Experts on Immunization (SAGE), vaccine hesitancy is the term used to describe: “delay in acceptance or refusal of vaccination despite availability of vaccination services” [1]. Factors that affect the attitude towards acceptance of vaccination include complacency, convenience and confidence [1,2]. Complacency denotes the low perception of the disease risk; hence, vaccination deemed unnecessary. Confidence refers to the trust in vaccination safety, effectiveness, besides the competence of the healthcare systems. Convenience entails the availability, affordability and delivery of vaccines in a comfortable context [2].

The complex nature of motives behind vaccine hesitancy can be analyzed using the epidemiologic triad of environmental, agent and host factors [3,4]. Environmental factors include public health policies, social factors and the messages spread by media [5-7]. The agent (vaccine and disease) factors involve the perception of vaccine safety and effectiveness besides the perceived susceptibility to the disease [7-9]. Host factors are dependent on knowledge, previous experience, educational and income levels [4,10].

The current coronavirus disease 2019 (COVID-19) pandemic does not seem to show any signs of decline, with more than 1.7 million deaths and more than 80 million reported cases worldwide, as of December 27, 2020 [11,12]. The ebb and flow of COVID-19 cases can be driven by human factors including attitude towards physical distancing and protective measures, while viral factors are driven by mutations that commonly occur in severe acute respiratory syndrome coronavirus 2 (SARS-COV-2) genome [13-18]. The viral factors can be particularly of high relevance considering the recent reports of resurgence in COVID-19 infections in UK due to a new variant of the virus [19].

The global efforts to lessen the effects of the pandemic, and to reduce the health and socio-economic impact relies to a large extent on the preventive efforts [20,21]. Thus, huge efforts by the scientific community and pharmaceutical industry backed by governments’ support, were directed towards developing efficacious and safe vaccines for SARS-CoV-2 [22]. These efforts were manifested by approval of three vaccines, in addition to more than 60 vaccine candidates in clinical trials. Moreover, more than 170 COVID-19 vaccine candidates are in the pre-clinical phase [23].

Despite the huge efforts made to achieve successful COVID-19 vaccines, a major hindrance can be related to vaccine hesitancy towards the approved and prospective COVID-19 vaccination [24]. To identify the scope of this problem, this systematic review aimed to assess the acceptance rates for COVID-19 vaccine(s) in different countries worldwide.

## Methods

### Eligibility criteria and search strategy

This review was conducted following the PRISMA guidelines [25].

Published papers in PubMed/Medline that aimed at evaluating COVID-19 vaccine hesitancy/vaccine acceptance using a survey/questionnaire were eligible for inclusion in this review.

Only studies in English language that met the inclusion criteria were considered in this review.

Search was done as of December 25, 2020 using the following strategy: (((COVID* vaccine* hesitancy[Title/Abstract]) OR (COVID* vaccine acceptance[Title/Abstract])) OR (COVID* vaccin* hesitanc*[Title/Abstract])) OR (COVID* intention to vaccin* [Title/Abstract]) OR (COVID* vaccin* accept*[Title/Abstract]) AND (2020:2020[pdat]).

Screening of titles and abstracts was conducted, followed by data extraction for the following items: Date of survey, country/countries in which the survey was conducted, target population for survey (e.g. general public, healthcare workers, students), total number of respondents, and COVID-19 vaccine acceptance rate (which included the number of respondents who answered: agree/somewhat/completely agree/leaning towards yes/definitely yes).

## Results

A total of 178 records were identified, and following the screening process, a total of 30 articles were included in this review (**Figure 1**). In addition, data collected in an unpublished manuscript that surveyed the general public residing in Jordan and Kuwait were added to the final analysis [26].

**Figure 1.**
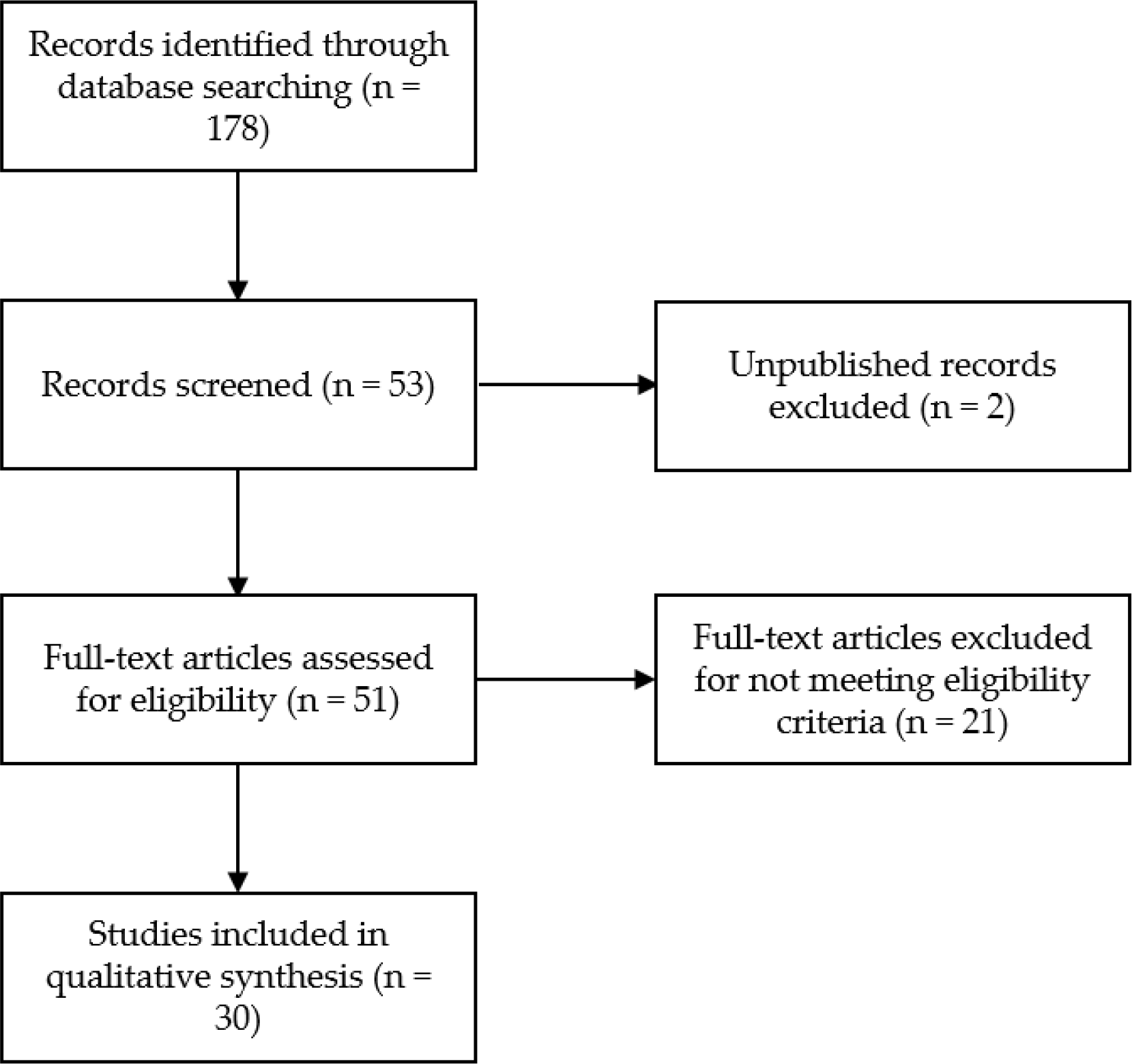
Flow chart of the study selection process.

### Characteristics of the papers included in this review

A total of 30 published papers were analyzed in this review, with an additional unpublished manuscript that focused on COVID-19 vaccine acceptance in Jordan and Kuwait to yield a total of 31 studies. These studies comprised surveys on COVID-19 vaccine acceptance from a total of 33 different countries. Surveys were done most commonly in UK (n=6), followed by France & US (n=5, for each country), and China & Italy (n=4, for each country). Dates of survey distribution ranged from February 2020 until December 2020. A few studies were conducted in more than one country; with the study by Lazarus *et al* involving 19 countries and the study by Neumann-Böhme *et al* involving seven European countries [27,28].

Stratified per country, a total of 60 surveys were found with the largest sample size (n=5114) in the study conducted in UK by Freeman *et al*, while the smallest sample size (n=123) was found in the study conducted in Malta by Gretch *et al* among general practitioners and trainees [29,30]. Out of these 60 surveys, 47 were among the general public, eight surveys were among healthcare workers (doctors, nurses, or others), three surveys were among parents/guardians and two surveys were among University students (**Table 1**). Surveys were most commonly conducted in June or July (23/60, 38%), followed by March or April (20/60, 33%).

**Table 1.** COVID-19 acceptance rates divided by the included studies and sorted based on date of survey.

| **Study** | **Country** | **Date of survey** | **N** | **Target population** | **Acceptance rate (%)** |
| --- | --- | --- | --- | --- | --- |
| Wang *et al* [31] | Hong Kong | February and March, 2020 | 806 | Nurses | 40.0 |
| Wang *et al* [32] | China | March, 2020 | 2058 | General population | 91.3 |
| Harapan *et al* [33] | Indonesia | March and April 2020 | 1359 | General population | 93.3 |
| Dror *et al* [34] | Israel | March and April 2020 | 388 | Doctors | 78.1 |
| Detoc *et al* [35] | France | March and April 2020 | 3259 | General population | 77.1 |
| Dror *et al* [34] | Israel | March and April 2020 | 1112 | General population | 75.0 |
| Kwok *et al* [36] | Hong Kong | March and April 2020 | 1205 | Nurses | 63.0 |
| Dror *et al* [34] | Israel | March and April 2020 | 211 | Nurses | 61.1 |
| Nzaji *et al* [37] | DRC | March and April 2020 | 613 | Healthcare workers | 27.7 |
| Gagneux-Brunon *et al* [38] | France | March to July, 2020 | 2047 | Healthcare workers | 75.9 |
| Sarasty *et al* [39] | Ecuador | April, 2020 | 1050 | General population | 97.0 |
| Wong *et al* [40] | Malaysia | April, 2020 | 1159 | General population | 94.3 |
| Neumann-Böhme *et al* [28] | Denmark | April, 2020 | 1000 | General population | 80.0 |
| Neumann-Böhme *et al* [28] | UK | April, 2020 | 1000 | General population | 79.0 |
| Neumann-Böhme *et al* [28] | Italy | April, 2020 | 1500 | General population | 77.3 |
| Ward *et al* [41] | France | April, 2020 | 5018 | General population | 76.0 |
| Neumann-Böhme *et al* [28] | Portugal | April, 2020 | 1000 | General population | 75.0 |
| Neumann-Böhme *et al* [28] | Netherland | April, 2020 | 1000 | General population | 73.0 |
| Neumann-Böhme *et al* [28] | Germany | April, 2020 | 1000 | General population | 70.0 |
| Neumann-Böhme<br>*et al* [28] | France | April, 2020 | 1000 | General<br>population | 62.0 |
| Fisher *et al* [42] | US | April, 2020 | 1003 | General<br>population | 56.9 |
| Salali & Uysal<br>[43] | UK | May, 2020 | 1088 | General<br>population | 86.0 |
| Lin *et al* [44] | China | May, 2020 | 3541 | General<br>population | 83.5 |
| Taylor *et al* [45] | Canada | May, 2020 | 1902 | General<br>population | 80.0 |
| Taylor *et al* [45] | US | May, 2020 | 1772 | General<br>population | 75.0 |
| Salali & Uysal<br>[43] | Turkey | May, 2020 | 3936 | General<br>population | 69.0 |
| Reiter *et al* [46] | US | May, 2020 | 2006 | General<br>population | 68.5 |
| Malik *et al* [47] | US | May, 2020 | 672 | General<br>population | 67.0 |
| Lazarus *et al* [27] | China | June, 2020 | 712 | General<br>population | 88.6 |
| Barello *et al* [48] | Italy | June, 2020 | 735 | University<br>students | 86.1 |
| Lazarus *et al* [27] | Brazil | June, 2020 | 717 | General<br>population | 85.4 |
| Lazarus *et al* [27] | South Africa | June, 2020 | 619 | General<br>population | 81.6 |
| Lazarus *et al* [27] | South Korea | June, 2020 | 752 | General<br>population | 79.8 |
| Lazarus *et al* [27] | Mexico | June, 2020 | 699 | General<br>population | 76.3 |
| Lazarus *et al* [27] | US | June, 2020 | 773 | General<br>population | 75.4 |
| Lazarus *et al* [27] | India | June, 2020 | 742 | General<br>population | 74.5 |
| Lazarus *et al* [27] | Spain | June, 2020 | 748 | General<br>population | 74.3 |
| Lazarus *et al* [27] | Ecuador | June, 2020 | 741 | General<br>population | 71.9 |
| Lazarus *et al* [27] | UK | June, 2020 | 768 | General<br>population | 71.5 |
| Lazarus *et al* [27] | Italy | June, 2020 | 736 | General<br>population | 70.8 |
| Lazarus *et al* [27] | Canada | June, 2020 | 707 | General<br>population | 68.7 |
| Lazarus *et al* [27] | Germany | June, 2020 | 722 | General population | 68.4 |
| Lazarus *et al* [27] | Singapore | June, 2020 | 655 | General population | 67.9 |
| Lazarus *et al* [27] | Sweden | June, 2020 | 650 | General population | 65.2 |
| Lazarus *et al* [27] | Nigeria | June, 2020 | 670 | General population | 65.2 |
| Lazarus *et al* [27] | France | June, 2020 | 669 | General population | 58.9 |
| Lazarus *et al* [27] | Poland | June, 2020 | 666 | General population | 56.3 |
| Lazarus *et al* [27] | Russia | June, 2020 | 680 | General population | 54.9 |
| Rhodes *et al* [49] | Australia | June, 2020 | 2018 | Parents and guardians | 77.3 |
| Bell *et al* [50] | UK | July, 2020 | 1252 | Parents and guardians | 90.1 |
| Sherman *et al* [51] | UK | July, 2020 | 1500 | General population | 64.0 |
| Zhang *et al* [52] | China | September, 2020 | 1052 | Parents and guardians | 72.6 |
| Gretch *et al* [30] | Malta | September, 2020 | 123 | GPs and GP trainees | 61.8 |
| La Vecchia *et al* [53] | Italy | September, 2020 | 1055 | General population | 53.7 |
| Gretch *et al* [54] | Malta | September, 2020 | 1002 | Healthcare workers | 52.0 |
| Gretch & Gauci [55] | Malta | September, 2020 | 852 | University students | 44.2 |
| Freeman *et al* [29] | UK | September and October, 2020 | 5114 | General population | 71.7 |
| Al-Mohaithef & Badhi [56] | Saudi Arabia | Unknown | 992 | General population | 64.7 |
| Sallam *et al* [26] | Jordan | December, 2020 | 2173 | General population | 28.4 |
| Sallam *et al* [26] | Kuwait | December, 2020 | 771 | General population | 23.6 |

### Rates of COVID-19 vaccine acceptance

The results of COVID-19 vaccine acceptance rates in different studies included in this review and stratified by country are shown in (**Table 1**). Classified per study, the highest vaccine acceptance rates (>90%) among the general public were found in four studies from Ecuador (97.0%), Malaysia (94.3%), Indonesia (93.3%) and China (91.3%). On the contrary, the lowest vaccine acceptance rates (<60%) among the general public were found in seven studies from Kuwait (23.6%), Jordan (28.4%), Italy (53.7), Russia (54.9%), Poland (56.3%), US (56.9%), and France (58.9%).

For the eight studies conducted among healthcare workers, three surveys reported vaccine acceptance rates below 60%, with the highest rate being among doctors in Israel (78.1%) and the lowest vaccine acceptance rate (27.7%) reported among healthcare workers in the Democratic Republic of the Congo (DRC).

For the three studies conducted among parents/guardians, the vaccine acceptance rates were more than 70%. For the two studies among University students, the vaccine acceptance rate was 44.2% in Malta and 86.1% in Italy.

### Changes in COVID-19 vaccine acceptance over time in countries with multiple survey studies

In countries with multiple surveys over time, the following changes in COVID-19 vaccine acceptance rates were observed: In UK, the vaccine acceptance rate was 79.0% in April, 86.0% in May, 71.5% in June, 64.0% in July and 71.7% in September/October. In France, the vaccine acceptance rate ranged from 62.0% to 77.1% in March/April and was 58.9% in June. In Italy, the vaccine acceptance rate was 77.3% in April, 70.8% in June and reached 53.7% in September.

For the vaccine acceptance rates in US, it was 56.9% in April, and ranged from 67.0% to 75.0% in May, and reached 75.4% in June. In China, three studies reported high rates of vaccine acceptance with the first study that reported a vaccine acceptance rate of 91.3% in March, the second study reported a rate of 83.5% in May and the third study reported a rate of 88.6% in June.

## Discussion

Vaccine hesitancy is an old phenomenon that represents a serious threat to the global health, as shown in resurgence of some infectious diseases (e.g. outbreaks of measles and pertussis) [57-61]. The huge leaps in developing efficacious and safe COVID-19 vaccines within a short period were unprecedented [62-64]. Nevertheless, COVID-19 vaccine hesitancy can be the limiting step in the global efforts to control the current pandemic with its negative health and socio-economic effects [24,65,66].

Assessing the level of population immunity necessary to limit the pathogen spread is dependent on the basic reproductive number for that infectious disease [67]. The latest estimates on COVID-19, pointed to the range of 60-75% immune individuals that would be necessary to halt the forward transmission of the virus and community spread of the virus [68-70]. Vaccine cost, effectiveness and duration of protection appear as important factors to achieve such a goal [64,71,72]. However, vaccine hesitancy can be a decisive factor for successful control of the current COVID-19 pandemic [24,73]. Thus, estimates of vaccine acceptance rates can be helpful to plan actions and intervention measures necessary to increase the awareness and assure people about the safety and benefits of vaccines, which in turn would help to control virus spread and alleviate the negative effects of this unprecedented pandemic [74,75].

In this review, a large variability in COVID-19 vaccine acceptance rates was found. However, certain patterns can be deduced based on descriptive analysis of the reported vaccine acceptance rates. First, in East and South East Asia, the overall acceptance rates among the general public were relatively high. This includes more than 90% acceptance rates in Indonesia, Malaysia and one study from China [32,33,40]. Another two surveys on the general public in China reported vaccine acceptance rates of more than 80%, with an additional survey in South Korea that reported a rate of 79.8% [27,44]. A later survey from Shenzhen, China by Zhang *et al*, that surveyed parents/guardians who were faculty workers, on their acceptability of children COVID-19 vaccination reported a lower rate of 72.5% compared to the previous studies [52]. Similarly, an online survey on Australian parents showed an acceptance rate of 77.3% [49]. The lowest COVID-19 vaccine acceptance rate among the general public in the region was reported by Lazarus *et al*, in Singapore (67.9%) [27]. The relatively high rates of vaccine acceptance in the region were attributed to strong trust in governments [27]. Additionally, the only survey in India reported a vaccine acceptance rate of 74.5% [27].

However, two studies that dated back to early part of the pandemic (February and March) among nurses in Hong Kong reported low rates of COVID-19 acceptance (40.0% and 63.0%) [31,36]. Likewise, Kabamba Nzaji *et al* reported a very low rate of COVID-19 vaccine acceptance among healthcare workers in the DRC (27.7%) [37]. This issue is of high concern considering the front-line position of healthcare workers in fighting the spread and effects of COVID-19 pandemic, which put them at a higher risk of infection, and hence their higher need for protective measures [76-78].

Also, the vaccine acceptance rates were relatively high in Latin America, where results from Brazil and Ecuador reported more than 70% acceptance rates [27,39]. This was also seen in the survey from Mexico with a vaccine acceptance rate of 76.3% [27].

In Europe, the results were largely variable, with countries around the Mediterranean reporting vaccine acceptance rates as low as 53.7% in Italy, 58.9% in France; while no surveys among general public in Malta were found [27,53]. However, the vaccine acceptance rates among students and healthcare workers in Malta were 44.2% and 52.0%, respectively [54,55]. Variable results were also reported in other European countries with rates as high as 80.0% in Denmark, and as low as 56.3% in Poland [27,28]. The vaccine acceptance rates were even lower in Russia (54.9%), which needs further evaluation considering the heavy toll of COVID-19 on the country [11,27]. Variability in vaccine acceptance rates was also seen in UK, US and Canada over the course of the pandemic [42,43,45,46,51].

The Middle East was among the regions with the lowest vaccine acceptance rates globally. The acceptance rate was the lowest in Kuwait (23.6%), Jordan (28.4%), Saudi Arabia (64.7%) and Turkey (69.0%) [26,43,56]. On the other hand, the highest vaccine acceptance rate was reported in Israel (75.0%), however; this rate was much lower among nurses surveyed in the same study (61.1%) [34].

Finally, only two surveys among the general public in African countries were found that reported an acceptance rate of 81.6% in South Africa and 65.2% in Nigeria [27]. Thus, more studies are recommended in Africa to address COVID-19 vaccine hesitancy in the continent. Besides Africa, more studies are needed from Central Asia, Eastern Europe, Central and South America to reach reliable conclusions about the scope of COVID-19 vaccine hesitancy around the globe.

## Conclusions

Large variability in COVID-19 vaccine acceptance rates was reported in different countries and regions of the world. A sizable number of studies reported COVID-19 acceptance rates below 60%, which would pose a serious problem for efforts to control the current COVID-19 pandemic. Low COVID-19 vaccine acceptance rates were more pronounced in the Middle East, Eastern Europe and Russia. High acceptance rates in East and South East Asia would help to achieve proper control of the pandemic. More studies are recommended to assess the attitude of general public and healthcare workers in Africa, Central Asia and the Middle East besides Central and South America. Such studies would help to evaluate COVID-19 vaccine hesitancy and its potential consequences in these regions, and around the globe.

## Data Availability

Data supporting this systematic review are available in the reference section. In addition, the analyzed data that were used during the current systematic review are available from the author on reasonable request.

